# The Association of Racial Residential Segregation and Survival After Out-of-Hospital Cardiac Arrest in the United States

**DOI:** 10.1101/2024.04.22.24306186

**Authors:** Ethan E Abbott, David G Buckler, Aditya C Shekhar, Elizabeth Landry, Benjamin S Abella, Lynne D Richardson, Alexis M Zebrowski, CARES Surveillance Group

**Author notes:** Corresponding Author: Ethan Abbott, DO, MSCR Address for Correspondence: Icahn School of Medicine at Mount Sinai Department of Emergency Medicine, 1 Gustave L. Levy Place, Box 1620 New York, NY 10029.

## Abstract

**Background:** Residential segregation has been identified as drivers of disparities in health outcomes, but further work is needed to understand this association with clinical outcomes for out-of-hospital cardiac arrest (OHCA). We utilized Cardiac Arrest Registry to Enhance Survival (CARES) dataset to examine if there are differences in survival to discharge and survival with good neurological outcome, as well as likelihood of bystander CPR, using validated measures of racial, ethnic, and economic segregation.

**Methods:** We conducted a retrospective observational study using data from the Cardiac Arrest Registry to Enhance Survival (CARES) dataset to examine associations among adult OHCA patients. The primary predictor was the Index of Concentration at the Extremes (ICE), a validated measure that includes race, ethnicity, and income across three measures at the census tract level. The primary outcomes were survival to discharge and survival with good neurological status. A multivariable modified Poisson regression modeling approach with random effects at the EMS agency and hospital level was utilized.

**Results:** We identified 626,264 OHCA patients during the study period. The mean age was 62 years old (SD 17.2 years), and 35.7% (n =223,839) of the patients were female. In multivariable models, we observed an increased likelihood of survival to discharge and survival with good neurological outcome for those patients residing in predominately White population census tracts and higher income census tracts as compared to lower income Black and Hispanic/Latinx population census tracts (RR 1.24, CI 1.20-1.28) and a 32% increased likelihood of receiving bystander CPR in higher income census tracts as compared to reference (RR 1.32, CI 1.30-1.34).

**Conclusions:** In this study examining the association of measures of residential segregation and OHCA outcomes, there was an increased likelihood of survival to discharge, survival with good neurological status, and likelihood of receiving B-CPR for those patients residing in predominately White population and higher income census tracts when compared to predominately Black and/or Hispanic Latinx populations and lower income census tracts. This research suggests that areas impacted by residential and economic segregation are important targets for both public policy interventions as well as addressing disparities in care across the chain of survival for OHCA.

## Introduction

Social determinants of health (SDOH) are recognized as important predictors for clinical outcomes across multiple health conditions, including out-of-hospital cardiac arrest (OHCA).^1–3^ Residential and economic segregation – an important SDOH domain and the result of historical institutional and systemic racism in the United States – has been tied to poor health outcomes across multiple chronic and acute health conditions.^4–9^ A sizeable body of research has demonstrated strong associations with OHCA clinical outcomes and several key SDOH factors. Neighborhood racial composition and socioeconomic status have been shown to be associated with differences in rates of bystander CPR, leading to disparities in survival to discharge and survival with good neurological status.^3,10–12^

In our prior OHCA research, we found that measures of racial and economic segregation are predictors of disparities in outcomes; Medicare beneficiaries residing in more highly segregated Black population and lower income ZIP codes had decreased likelihood of survival to discharge and survival at one year, as well as increased risk of readmission at 30 days.^13, 14^ However, prehospital predictors are not available in claims data, such as initial rhythm or the presence of a bystander CPR (B-CPR). To better understand the association of measures of residential segregation with OHCA clinical outcomes while accounting for important prehospital variables, we utilized Cardiac Arrest Registry to Enhance Survival (CARES) dataset to examine if there are differences in survival to discharge and survival with good neurological outcome, as well as likelihood of bystander CPR, using validated measures of racial, ethnic, and economic segregation.

## Methods

### Study Design and Patient Population: The Cardiac Arrest Registry to Enhance Survival (CARES)

We conducted a retrospective observational study using data from the Cardiac Arrest Registry to Enhance Survival (CARES) between January 1, 2013, to December 31, 2022. For this analysis, we included adult patients (>18 years old) and initially attended by a non-EMS bystander. We utilized a threshold for EMS agencies with a minimum of 10 OHCA events across the study period. Exclusions for the cohort were any OHCAs witnessed by a 911 responder, those at healthcare facilities, or if the primary predictor variables were missing (Figure 1). CARES is a federally supported registry aimed at improving data collection for OHCA and specific details about the registry have been published previously.^15, 16^ Briefly, participating centers complete standardized data elements about out-of-hospital cardiac arrest cases, including prehospital and in-hospital variables. Cases are then reviewed for completeness and accuracy by research coordinators before being submitted. The registry follows the Utstein template for data collection in cardiac arrest, which has been supported by multiple international societies.^17, 18^ Currently, more than 2,300 EMS agencies and 2,500 hospitals participate in CARES data collection, representing a catchment area of approximately 179 million – which includes 34 statewide registries.

**Figure 1:**
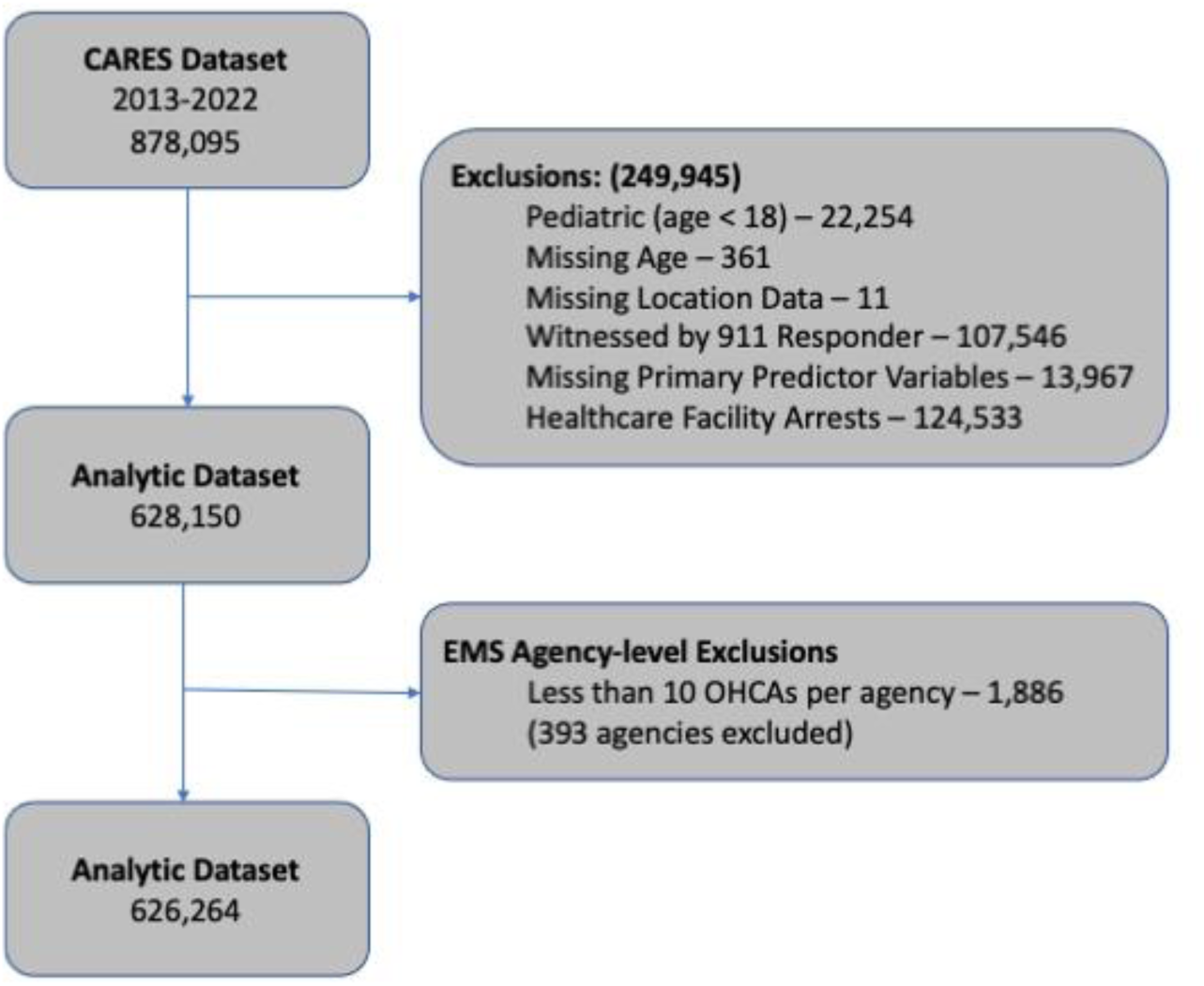
Study flow Diagram for OHCA CARES Cohort 2013-2022.

### ICE Measures

The primary predictor for this study was the Index of Concentration at the Extremes (ICE), a validated measure for assessing segregation based on race, ethnicity, income, or a combination of both. This methodology has been previously applied in our research exploring both short and long-term outcomes in OHCA.^13, 14^ The ICE measure is considered a more methodologically robust method as compared with other measures such as the usage of racial of ethnic percentage composition of a given geographic area (census tract, county, ZIP code). ICE captures geographies where extreme differences in race, ethnicity, and economic segregation exist—factors that are potential drivers of disparities in health outcomes.^19, 20^

The ICE measure is derived by calculating the difference between the numbers of individuals in privileged and underprivileged groups, and then dividing this by the overall population count (supplemental Figure 1 for operationalized formula). Essentially, the index represents the proportional difference between more advantaged and less advantaged groups. Drawing from our previous research utilizing the ICE framework, we computed three distinct indices: 1) ICE race and/or ethnicity 2) ICE income 3) ICE race (and/or ethnicity) + income (a combined measure). The data for these calculations were sourced from the American Community Survey five-year estimates by the US Census Bureau, using census tract as the geographic unit of analysis. For all the ICE measures, we focused on Black, Hispanic/Latinx, and White populations because these groups represent the extremes of racial privilege and disadvantage in the US and the dataset did not include adequate sample sizes for a robust analysis of all other racial and ethnic categories collected by CARES. We categorized the lowest income bracket as under $25,000 and the highest as $100,000 or more, reflecting the 20th and 80th income percentiles in the US, a standardized approach when using ICE. The ICE indices are continuous, ranging from −1 (indicating maximum deprivation) to 1 (indicating maximum privilege). To enhance the usability and clarity of these measures, we categorized the ICE scores for each census tract into quintiles. Quintile 5 (Q5) denotes census tracts with the highest economic advantage, while quintile 1 (Q1) indicates those with the least. For the combined race and income ICE measures, our analysis utilized high-income White individuals (earning ≥$100,000) with low-income Black or Hispanic or Latinx individuals (earning <$25,000), following established methodology for using these measures. We determined the ICE for each census tract by subtracting the count of high-income White persons from that of low-income Black persons or Hispanic/Latinx persons, relative to the total population in that census tract. These calculations were then categorized at the patient level.

### Covariates of Interest

Our covariates of interest included demographic characteristics including age, sex, race, and ethnicity. We also included key arrest characteristics, including if the arrest was witnessed, shockable vs. nonshockable first rhythm, presence of B-CPR, and arrest location.

### Outcomes

The primary outcomes were survival to discharge and survival with good neurological status as measured by Cerebral Performance Category (CPC) of 1. A CPC score of 1 is representative of individuals having no significant functional or cognitive disability. Our secondary outcome was the likelihood of receiving B-CPR by ICE measure and quintile. We also performed a sensitivity analysis to determine if there were average differences in the likelihood of an individual receiving B-CPR or if an AED was applied across all ICE quintiles and measures.

### Statistical Analysis

For descriptive statistics, we calculated means with standard deviation or medians with interquartile ranges for continuous covariates of interest and frequencies with proportions for nominal variables. For the primary predictor, we utilized three measures to test associations with clinical outcomes: 1) ICE race and/or ethnicity measure 2) ICE income measure (which includes all races and ethnicities), and an ICE measure that includes both race or ethnicity and income (race and ethnicity + income). For all analyses, Q1 represented the reference category.

We first determined relative risk (RR) of survival to discharge, survival with good neurological outcome, and likelihood of B-CPR for each ICE measure in univariate. We then utilized a multivariable modified Poisson regression modeling approach with random effects at the EMS agency and hospital level for the outcomes of survival to discharge and survival with good neurological status. For these multivariable models, we controlled for age, race, ethnicity, arrest witness status, shockable vs. non-shockable rhythm, presence or absence of B-CPR, and public location. To ensure the robustness of our results, we conducted a sensitivity analysis to examine if there were differences in average automated external defibrillator (AED) usage and B-CPR by ICE measure and quintile, controlling for random effects at the EMS agency level. This study was completed in accordance with the STROBE guidelines. The study was approved by Icahn School of Medicine at Mount Sinai Institutional Review Board (Study #23-00195). Data is available through submission of a proposal and completion of a data usage agreement with CARES.

## Results

### Overall Cohort

After exclusions, we identified 626,264 patients who suffered an OHCA during the study period treated by a total of 1691 EMS agencies. The mean age was 62 years old (SD 17.2 years) and 35.7% (n=223,839) of the patients were female (Table 1). By race and ethnicity, 125,959 were Black patients, 46,708 were Hispanic/Latinx patients, 313,465 were White patients, and 140,762 were Other/Unknown race or ethnicity patients. For first monitored rhythm, 80.1% (n=501,671) were classified as nonshockable, 16.8% (n=105,424) were in a public location, and 44.2% (n=277042) were witnessed by bystanders. Bystanders performed CPR in 39.6% (n=248179) of OHCAs and an AED was applied in 1.8% (11097) of these events. Overall survival to discharge for the cohort was 9.3% (n=58,368) and survival with good neurological outcome overall (CPC=1) was 6.0% (n=37,492). For ICE measures, overall distributions by census tract for each measure and quintile are provided in Supplemental Table 1.

**Table 1:**
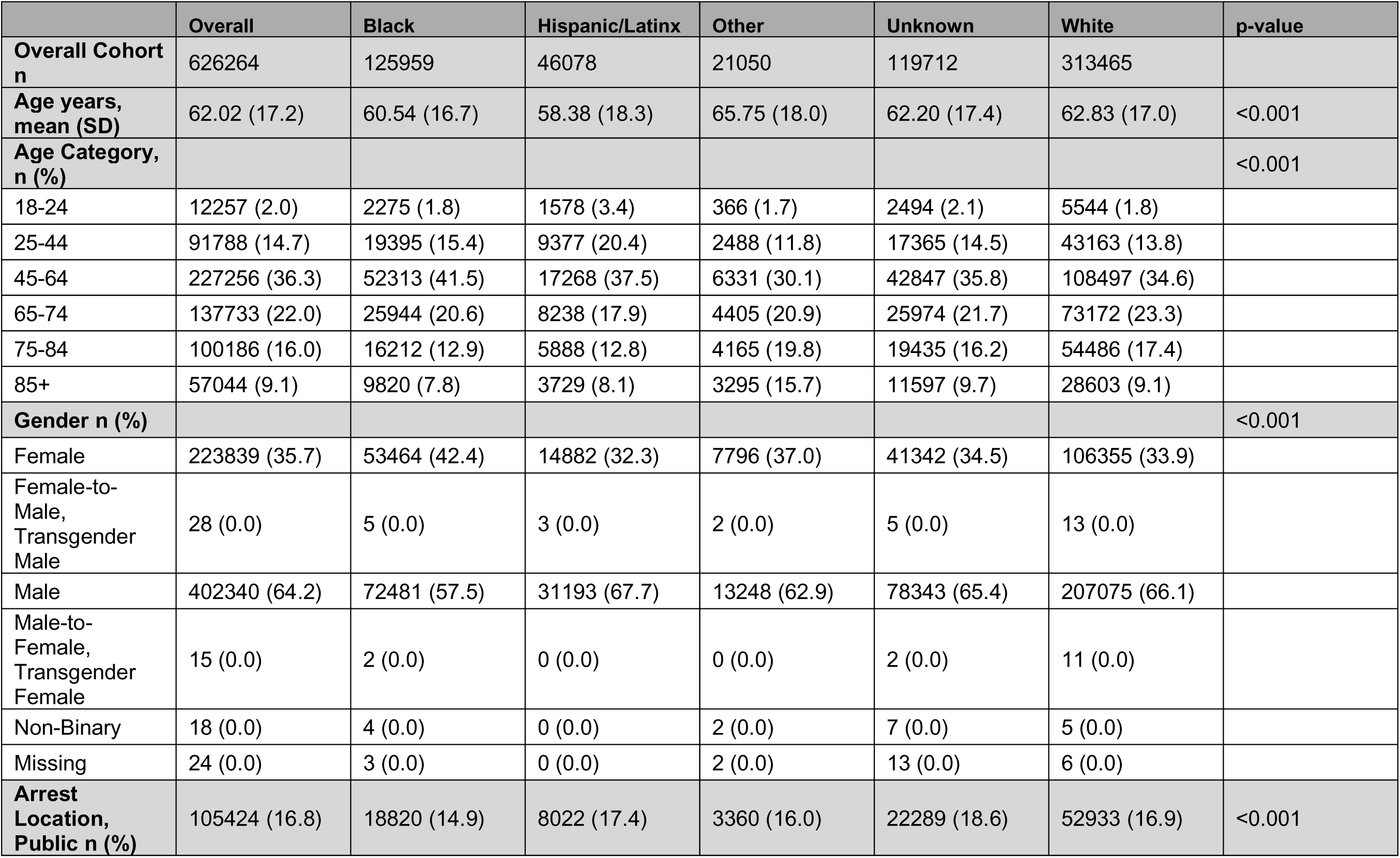
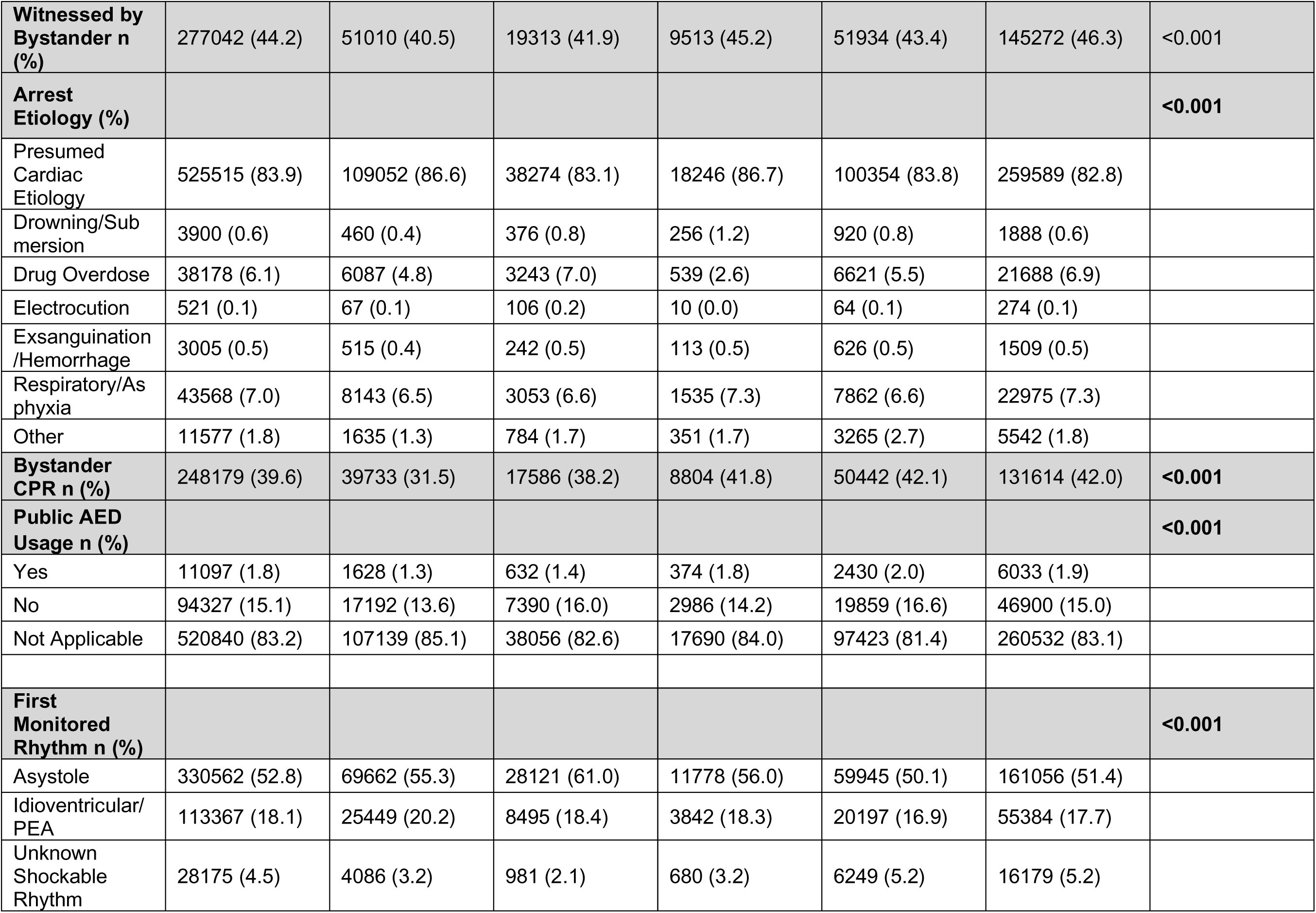
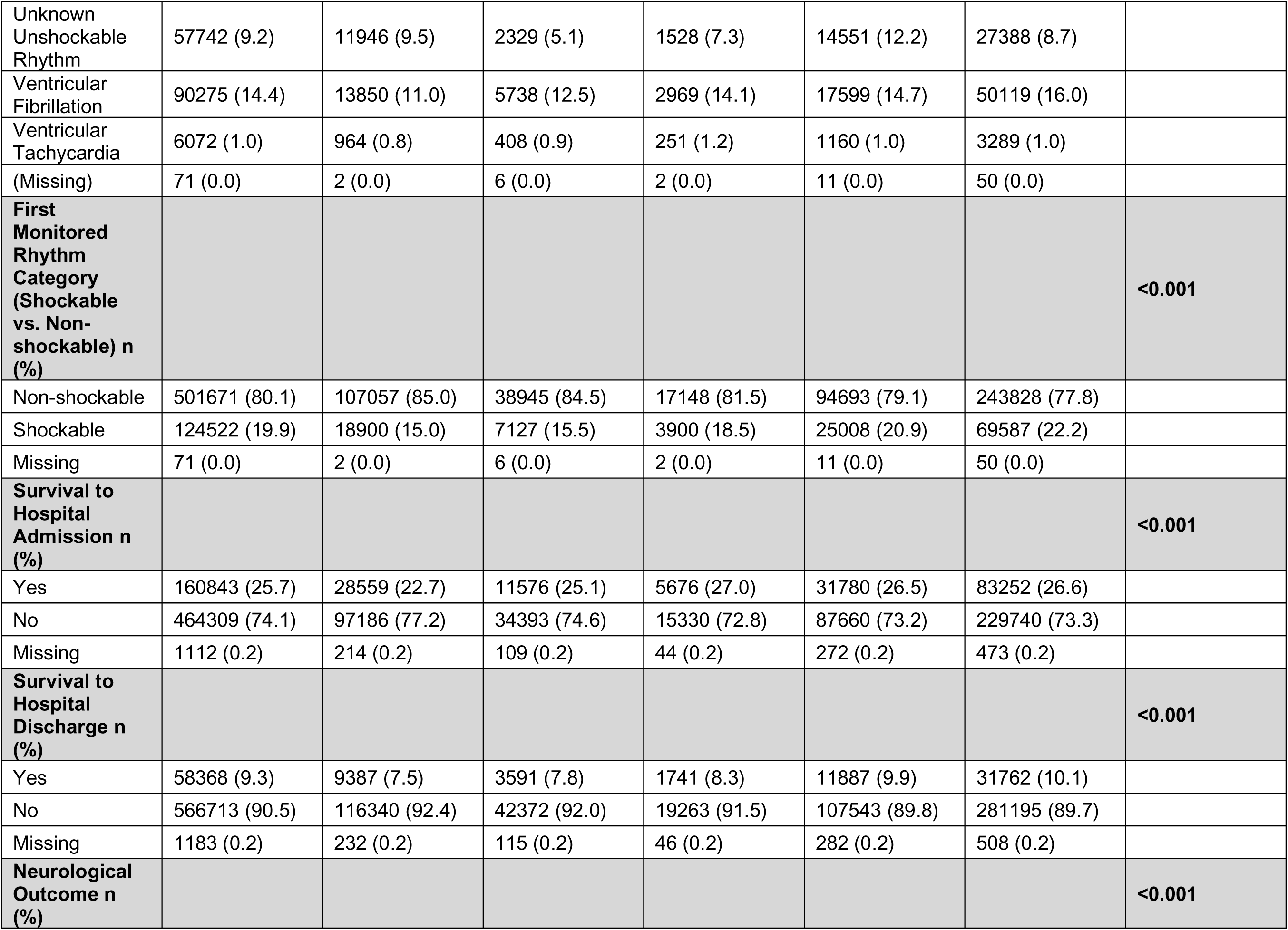
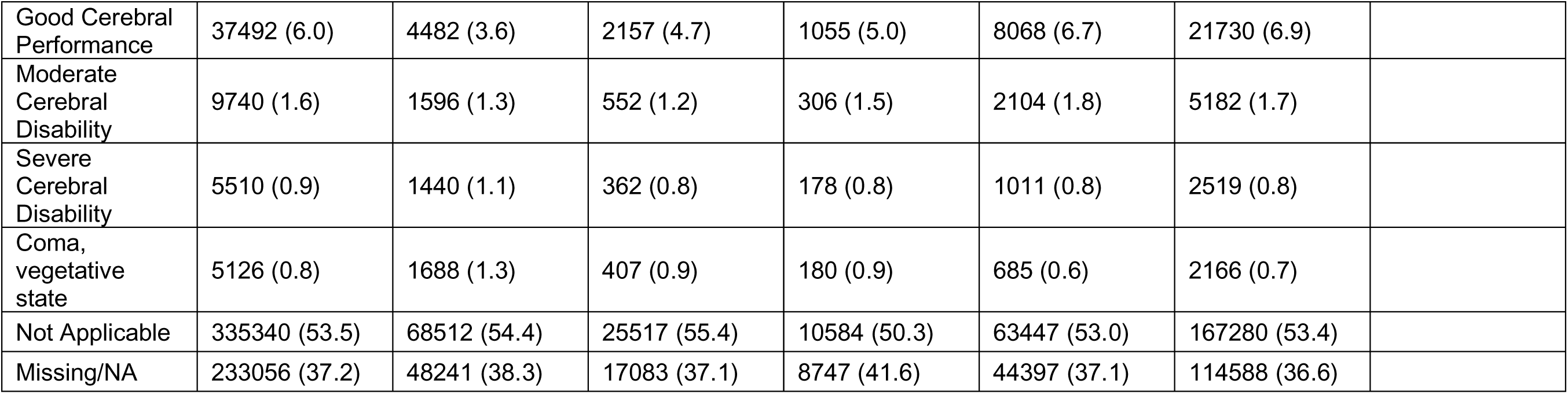
Overall Cohort Characteristics.

### Outcome: Survival to Discharge

In multivariable models controlling for age, sex, race and/or ethnicity, presence/absence of bystander witness, shockable vs. non-shockable, B-CPR, and arrest location, we found, for ICE measures that included Black and White patients, an increased likelihood of survival to discharge for those residing in more highly segregated predominately White populations and higher income census tracts (ICE Race Q5 RR 1.08, CI 1.04-1.12, ICE Race + Income RR 1.09, CI 1.05-1.12, Figure 2) as compared to reference. The likelihood of survival decreased from higher quintiles (Q5) to lower quintiles (Q1) when compared to reference and was statistically significant across all three measures. Shockable rhythm was the strongest independent predictor with an increased likelihood of survival to discharge (RR 2.07, CI 2.04-2.11). For multivariable models with ICE Income measure (all races and ethnicities) we found those patients residing in higher income census tracts had at least 7% increased likelihood of survival to discharge compared to lower income census tracts (ICE Income Measure Q5 RR 1.07, CI 1.04-1.10).

**Figure 2:**
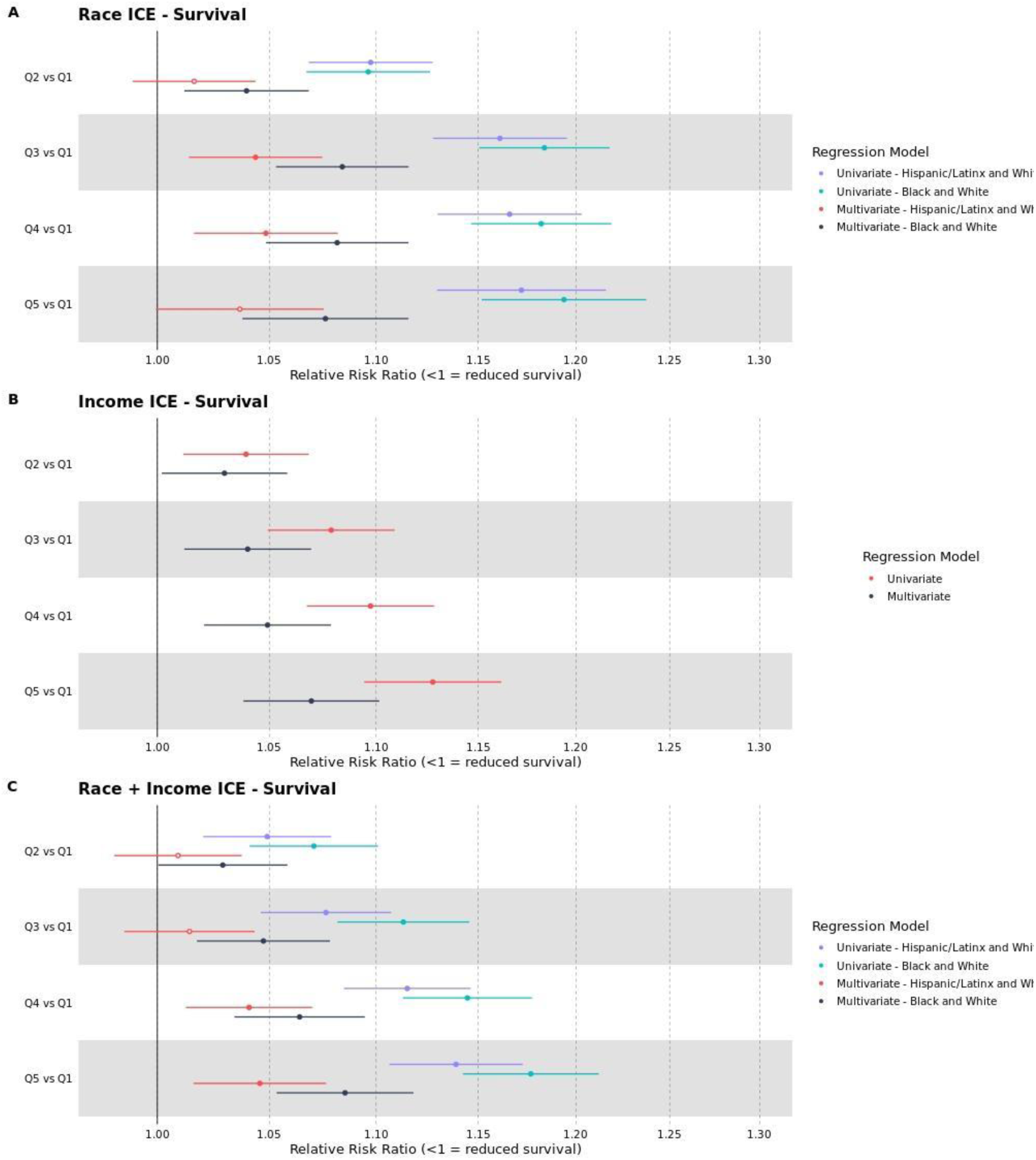
Univariable and Multivariable Models by ICE Measure for Black, Hispanic/Latinx, and White Patients for the Outcome of Survival to Discharge. Forest Plots of Univariate and Multivariable Models for the outcome of survival to discharge for all three ICE measures. For ICE race and ethnicity models, Q5 represents more highly segregated predominately White population census tracts, Q1 (reference) represents more highly segregated Black population or Hispanic/Latinx census tracts. For ICE Income Models Q5 represents higher income census tracts and Q1 represents lower income census tracts.

In multivariable models assessing Hispanic/Latinx and White patients, we found similar trends with an increased likelihood of survival to discharge for those patients residing in more highly segregated White populations and higher income census tracts as compared to more highly segregated Hispanic/Latinx populations and lower income census tracts (ICE Race Measure Q5 RR 1.04, CI 0.99-1.08, ICE Income Measure ICE Race and Ethnicity + Income Q5 1.00, CI 0.86-1.17) (Figure 2), however, this was not statistically significant. We also noted for the ICE Race + Income measure, there was a decreased likelihood of survival across all quintiles, but this also was not statistically significant.

### Outcome: Survival with Good Neurological Outcome

For the outcome of survival with good neurological status among Black and White patients, we found similar trends across all three measures and quintiles, but this association was notably more amplified by the ICE Race measure, as compared to other measure (ICE Race Measure Q5 RR 1.20, CI 1.15-1.24, ICE Race and Ethnicity + Income Measure Q5 RR 1.17, CI 1.14-1.22, Figure 3). Shockable rhythm was also the strongest independent predictor of survival with good neurological status (RR 2.35, CI 2.31-2.40). For the ICE Income Measure, we found an increased likelihood of survival for higher income quintiles as compared to reference (ICE Income Measure Q5 RR 1.13, CI 1.09-1.16, Figure 3). For the outcome of survival with good neurological status (CPC 1) among Hispanic/Latinx and White patients we found similar trends with increased likelihood of survival with good neurological outcome for those patients residing in Q5 (ICE Race Measure Q5 RR 1.16, CI 1.11-1.21, ICE Race + Income Measure Q5 RR 1.13, CI 0.91-1.39) however this was not statistically significant across all quintiles or measures (Figure 3).

**Figure 3:**
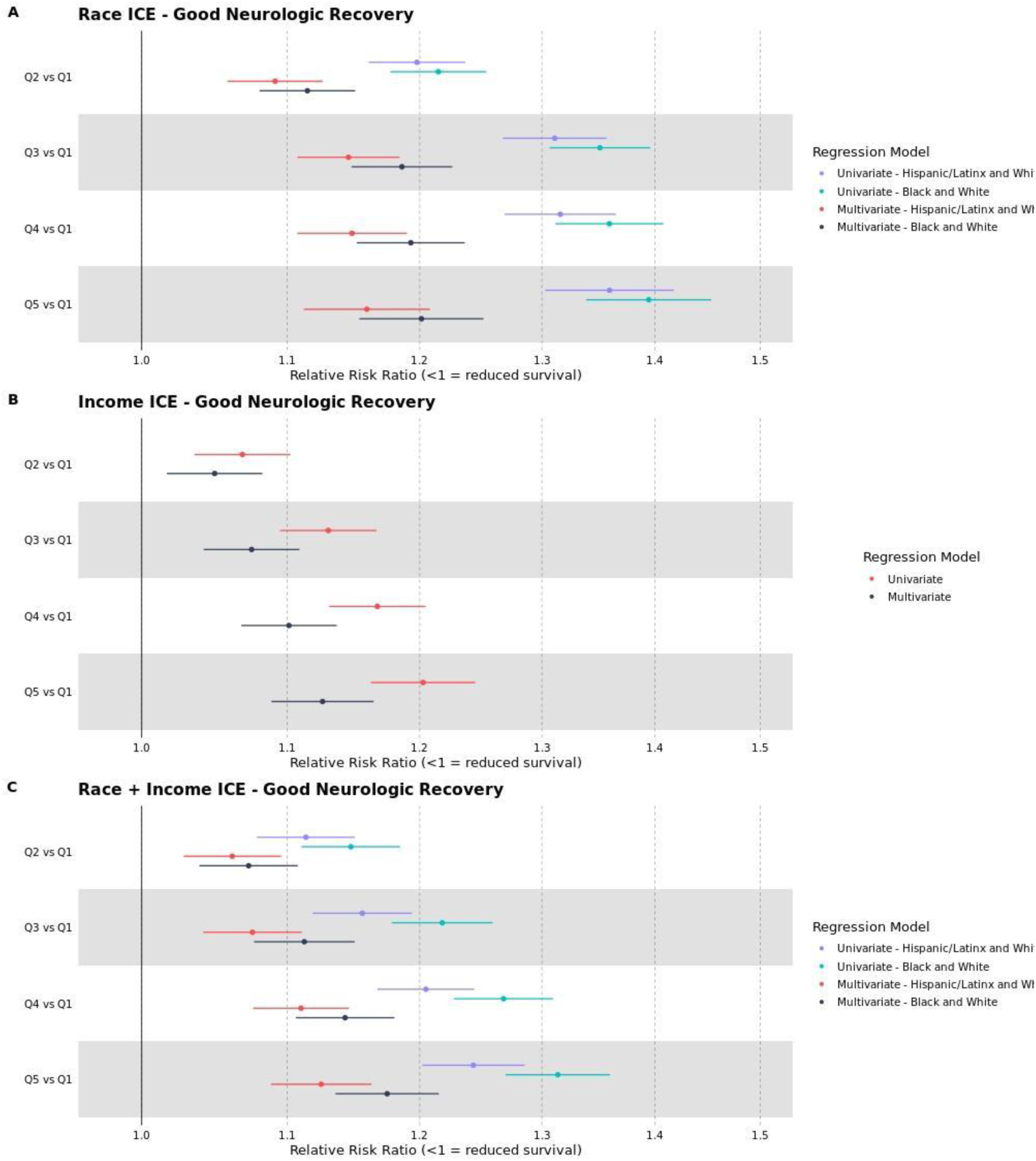
Univariable and Multivariable Models by ICE Measure for Black, Hispanic/Latinx, and White Patients for the Outcome of Survival with Good Neurological Outcome. Forest Plots of Univariate and Multivariable Models for the outcome of good neurological outcome (CPC 1) for all ICE measures. For ICE race and ethnicity models, Q5 represents more highly segregated predominately White population census tracts, Q1 (reference) represents more highly segregated Black population or Hispanic/Latinx census tracts. For ICE Income Models Q5 represents higher income census tracts and Q1 represents lower income census tracts.

### Outcome: Likelihood of B-CPR by ICE Measure

In our analysis of the secondary outcome of likelihood of receiving B-CPR by ICE measure and quintile, the ICE income measure was the strongest predictor; for those patients residing in Q5, the likelihood of receiving bystander CPR was 32% higher compared to reference (Q5: RR 1.32, CI 1.30-1.34, Figure 4). We also found similar patterns ICE measures that evaluated race, ethnicity, and combined measures. There was an increasing likelihood of a bystander performing CPR for those patients residing in more highly segregated, predominately White population and higher income census tracts compared to those residing in more highly segregated predominately Black population (ICE Race Q5 RR 1.30, CI 1.28-1.33) or Hispanic/Latinx population (ICE Race and Ethnicity Q5 RR 1.31, CI 1.28-1.33) census tracts.

**Figure 4:**
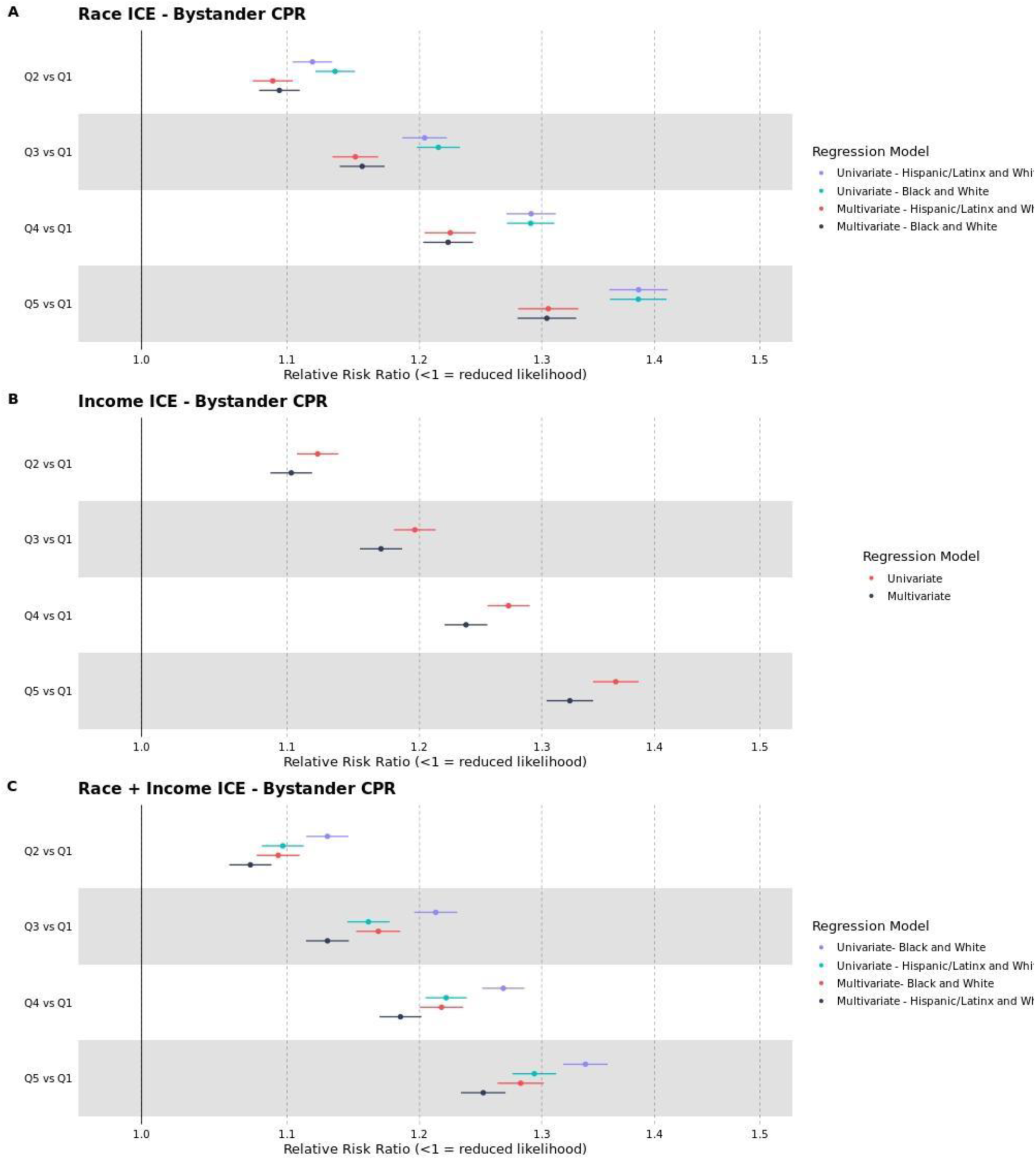
Univariable and Multivariable Models by ICE Measure for Black, Hispanic/Latinx, and White Patients for the Outcome of Likelihood of Receiving B-CPR. Forest Plots of Univariate and Multivariable Models for the outcome of likelihood of receiving B-CPR by ICE measure and quintile. For ICE race and ethnicity models, Q5 represents more highly segregated predominately White population census tracts, Q1 (reference) represents more highly segregated Black population or Hispanic/Latinx census tracts. For ICE Income Models Q5 represents higher income census tracts and Q1 represents lower income census tracts.

### Sensitivity Analysis

In a sensitivity analysis examining average B-CPR by quintile across all three measures there was a decreasing trend from Q5 to Q1 that was highest in the ICE Race + Income Measure for Black and White Patients (45.7%), and lowest for the same measure in Q1 (31.2%). For average AED usage by quintile and measure, the highest average AED usage was found in the ICE Race and Ethnicity + Income Measure for Hispanic/Latinx and White Patients in Q5 (13.1%) and lowest for the ICE Race and Ethnicity Measure for Hispanic/Latinx and White patients in Q1 (7.8%). The income measure and combined race and/or ethnicity measures trended similarly similar with average AED usage increasing linearly from Q1 to Q5. However, for the ICE Race measure we noted a decreasing trend from Q3 to Q5 for average AED usage for the ICE measures including both race and/or ethnicity and income (supplemental figure 3).

## Discussion

In this retrospective analysis of OHCA patients using CARES registry data, we observed an increased likelihood of survival to discharge and survival with good neurological outcome for those patients residing in more highly segregated predominantly White population and higher income census tracts as compared to more highly segregated and lower income Black and Hispanic/Latinx population census tracts using validated measures of racial, ethnic, and economic segregation. However, this finding was not statistically significant for patients residing in more highly segregated and lower income Hispanic/Latinx population census tracts. We also found a decreasing likelihood of B-CPR being performed in lower income and more highly segregated Black and Hispanic/Latinx census tracts across all three measures and quintiles. The likelihood of receiving B-CPR was 32% higher for populations residing in higher income census tracts. These results build on our prior research work examining Medicare beneficiaries who suffered an OHCA and reaffirms that key SDOH factors such as residential segregation are potential drivers of disparities in clinical outcomes and require further investigation and efforts to address health equity.

An emerging body of research has identified both broader area level SDOH domains as well as individual level health-related social needs (HRSN) as important non-clinical drivers of health outcomes. Studies of other emergent conditions, such as sepsis,^21, 22^ heart failure,^23^ and acute coronary syndrome,^24^ have shown that SDOH and HRSN data can be important for identifying drivers of disparities in outcomes and improving prediction specific clinical endpoints for patients. Specific to OHCA, a study examining 22,816 patients contained within the Resuscitation Outcomes Consortium (ROC) database found predominantly neighborhoods with a greater proportion of Black individuals were associated with lower rates of survival to hospital discharge when compared with neighborhoods with a greater proportion of White individuals.^25^ Similarly, a study examining 150,003 out-of-hospital cardiac arrests from the National Emergency Medical Services Information System (NEMSIS) found a relationship between neighborhood poverty level and a decreased likelihood of achieving return of spontaneous circulation (ROSC).^26^ And in a recent landmark study, also using CARES data, the authors compared the incidence of B-CPR in Black and Hispanic persons to White persons with consideration for racial, ethnic, and income composition at the neighborhood level. They found that Black and Hispanic persons were less likely to receive B-CPR at home or in public environments across all income strata and regardless of racial or ethnic neighborhood composition.^27^ In our study, we also found that at the census tract level, residential and economic segregation are associated with disparities in both clinical of our outcomes, but also impacting the likelihood of receiving B-CPR.

While public policy should work towards the end-goal of economic and structural revitalization of communities impacted by long-standing systemic racism resulting in residential and economic segregation, approaches to specifically address OHCA disparities based on these geographies will require an enhanced and renewed focus. This work should include actionable efforts targeted at delivering no cost or low-cost CPR training across impacted communities, advancing prehospital provider care and training, and a renewed focus on identifying mechanisms leading to disparities in OHCA outcomes among those surviving to hospital admission. To accomplish this, partnerships between community organizations, health care systems, and EMS agencies specific to census tracts, ZIP codes, or counties impacted by residential segregation should be the targets of these efforts. While complex, potential solutions will require improved data collection of non-clinical SDOH or HRSN factors and the ability to create linkages to OHCA clinical datasets to better understand the mechanisms leading to disparities in outcomes.

## Limitations

There are several important limitations to our study. Because the ICE is an area level measure and not individual level, there is potential for bias when examining associations with individual level health outcomes. Additionally, because of missing variables, we excluded more than 13,000 patients with missing ICE data which may have impacted our results. We also excluded patients who were transported by agencies with less than 10 OHCA per year, potentially excluding patients transported from rural or less populated census tracts, limiting the generalizability of our findings across all US populations. Despite these limitations, we believe that the methodology and conclusions from this work are robust and suggest that there are important disparities in OHCA outcomes to be addressed by communities impacted by long-standing residential and economic segregation.

## Conclusions

In this study examining the association of measures of residential segregation and OHCA outcomes using CARES data that included layperson witnessed arrests, we found an increased likelihood of survival to discharge and survival with good neurological status for those patients residing in predominantly White population and higher income census tracts when compared to reference. We also found similar trends in the likelihood of receiving B-CPR. These results build on our prior research that identified disparities in outcomes for OHCA Medicare beneficiaries across both short and long-term outcomes, suggesting that areas of the US impacted by segregation should be further targets for advancing care across the chain of survival for OHCA.

## Data Availability

All data produced in the present work are contained in the manuscript

## Acknowledgements

This work was supported in part through the computational and data resources and staff expertise provided by Scientific Computing and Data at the Icahn School of Medicine at Mount Sinai and supported by the Clinical and Translational Science Award (CTSA) grant UL1TR004419 from the National Center for Advancing Translational Sciences.

## Supplemental Tables and Figures

**Figure S1:**
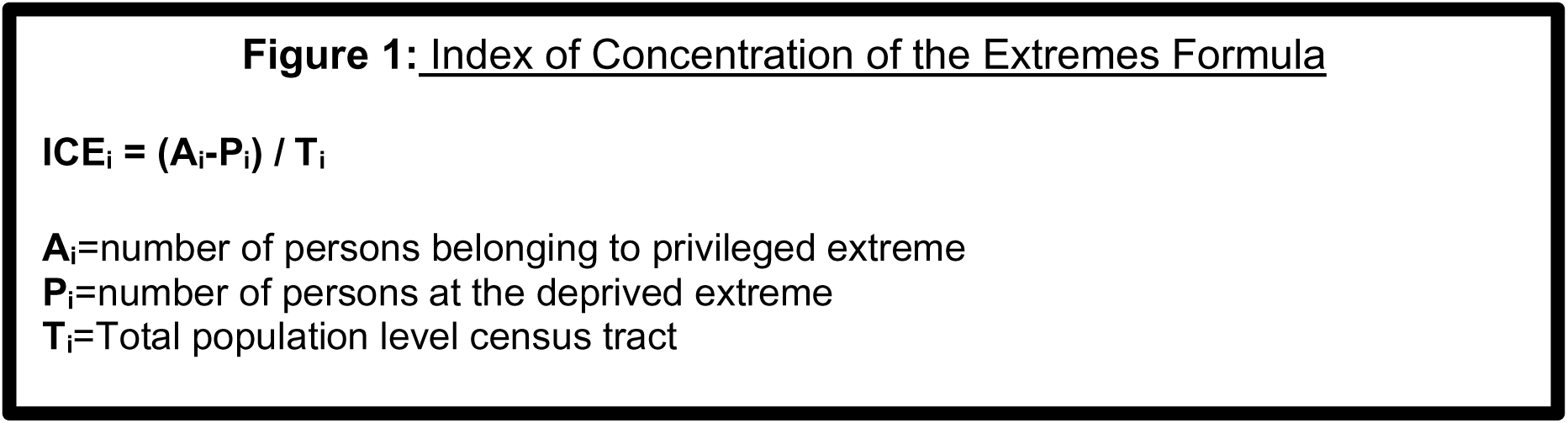
Index of Concentration of the Extremes (ICE) Operationalized Formula. The ICE measure is calculated as the difference between the count of privileged population and disadvantaged population, divided by the total population (census tract).

**Figure S2:**
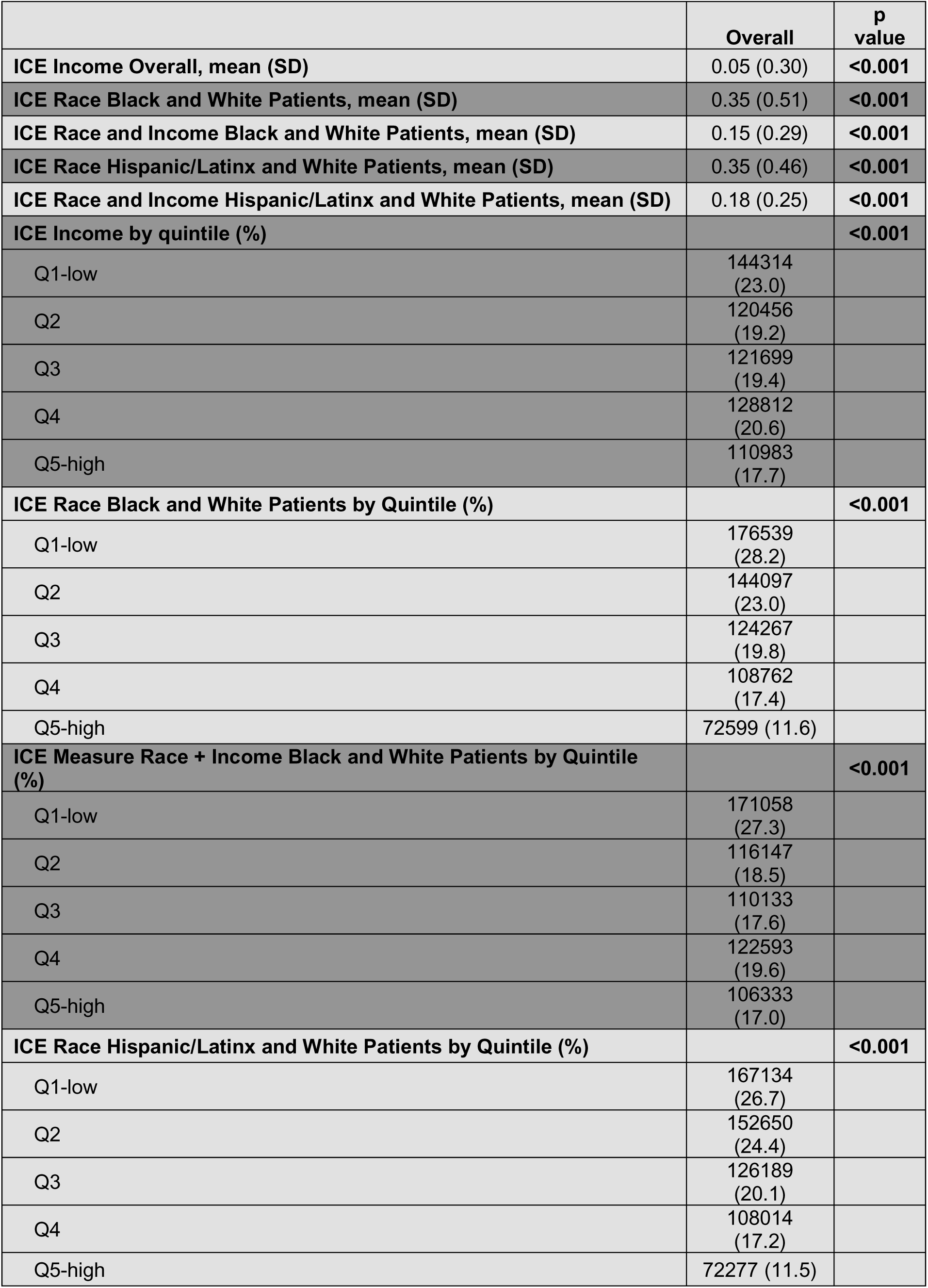
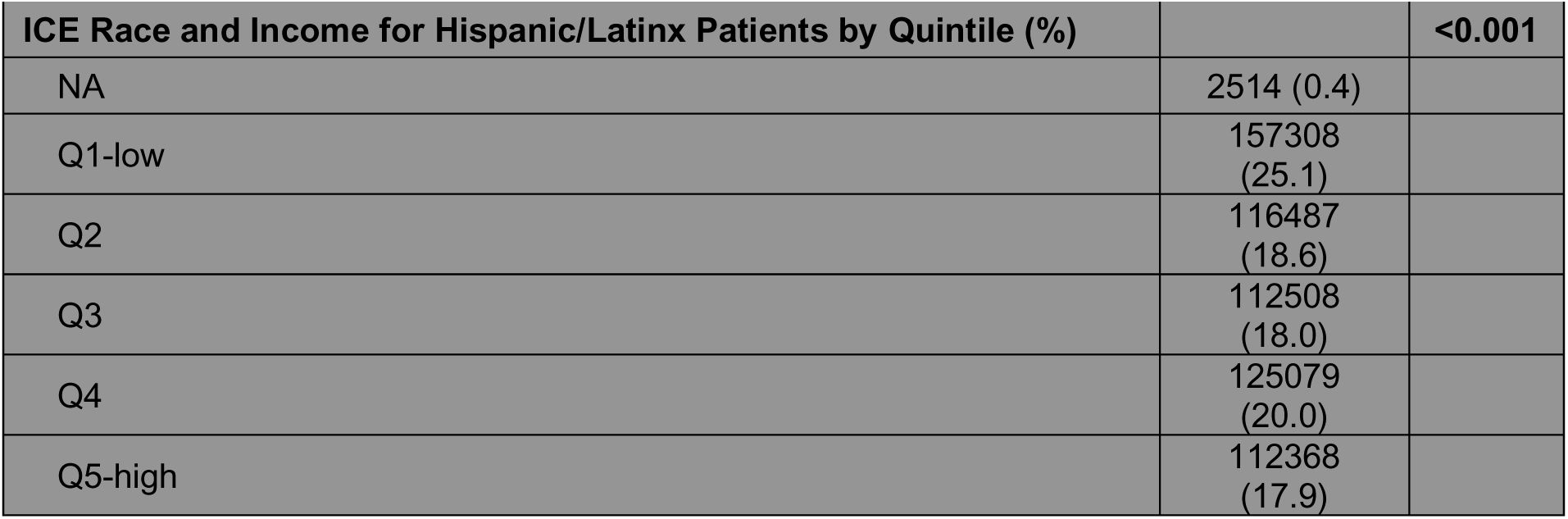
Raw ICE measures for census tract

**Figure S3:**
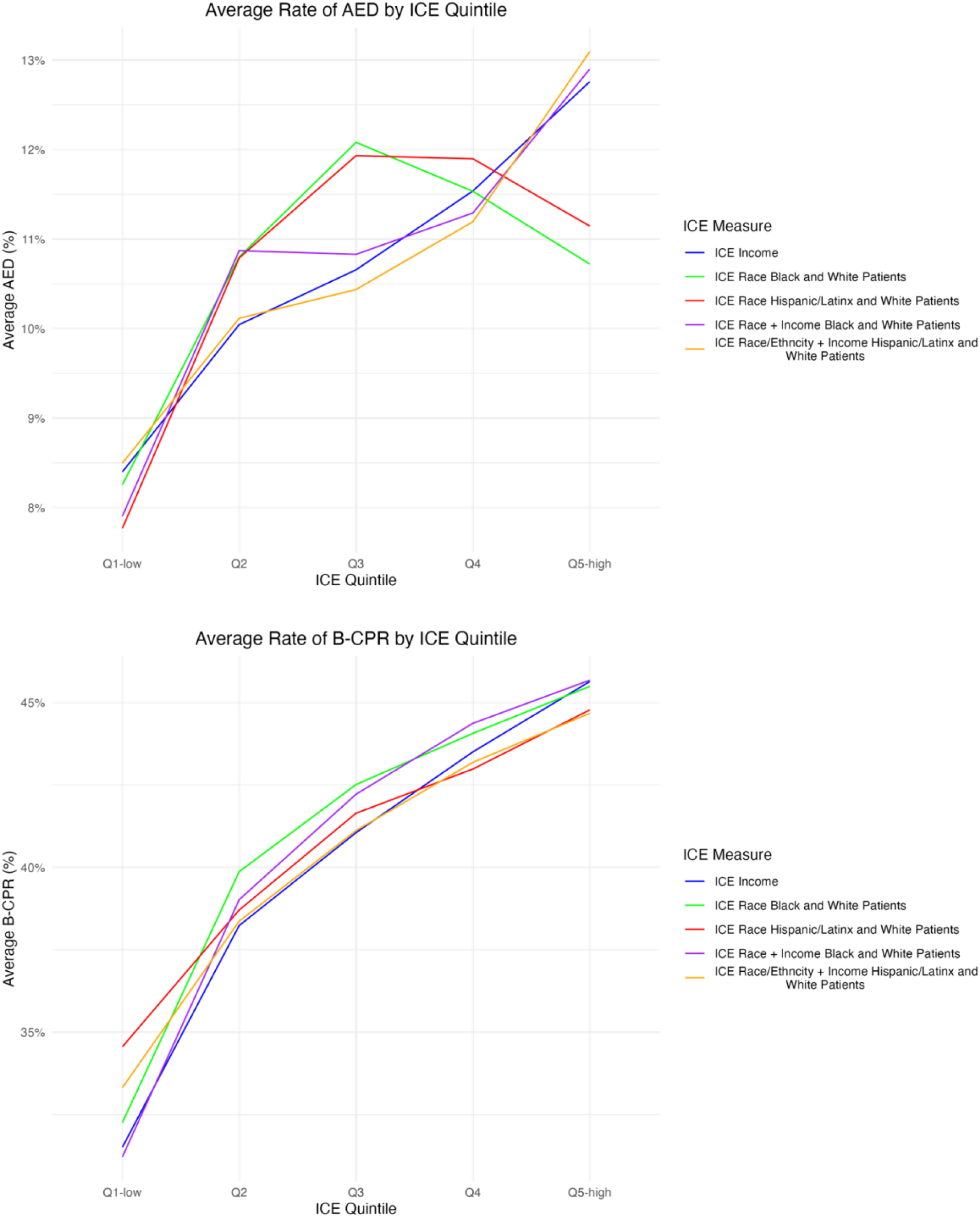
Sensitivity Analysis of Average Percentage Bystander Cardiopulmonary Resuscitation (B-CPR) and Average Automated External Defibrillator (AED) usage by ICE Measure and Quintile.

